# Characterization of ventricular tachycardia ablation in end-stage heart failure patients with left ventricular assist device (CHANNELED registry)

**DOI:** 10.1101/2024.10.30.24316462

**Authors:** J.-H. van den Bruck, F. Hohendanner, E. Heil, K. Albert, D. Duncker, H. Estner, T. Deneke, A. Parwani, E. Potapov, K. Seuthe, J. Wörmann, A. Sultan, J.-H. Schipper, L. Eckardt, F. Doldi, P. Lugenbiel, H. Servatius, G. Thalmann, T. Reichlin, M. Khalaph, D. Guckel, P. Sommer, D. Steven, J. Lüker

## Abstract

**Background:** Patients with left-ventricular-assist-devices (LVAD) are at high risk for ventricular tachycardia (VT), and data on VT ablation in LVAD patients is scarce. This multicenter registry assessed the mechanism of VT, procedural parameters, and outcome of VT ablation in LVAD patients (**NCT06063811)**.

**Methods:** Data of LVAD patients referred for VT ablation at 9 tertiary care centers were collected retrospectively. Parameters included VT mechanisms, procedural data, VT recurrence, and mortality.

**Results:** Overall, 69 patients (90% male, mean age 60.7±8.4 years) undergoing 72 catheter ablation procedures were included. Most procedures were conducted after intensification of antiarrhythmic drug (AAD) treatment (18/72; 25%) or after prior combination of ≥ 2 AADs (31/72; 43%). Endocardial low voltage areas were detected in all patients. 96 different VTs were targeted. The predominant mechanism was scar-related re-entry (76/96 VTs; 79%) and 19/96 VTs (20%) were related to the LVAD cannula. Non-inducibility of any VT was achieved in 28/72 procedures (39%). No LVAD related complication was observed. The extent of endocardial scar was associated with VT recurrence. Over a median follow-up of 283 days (IQR 70-587 days), 3/69 were lost to follow-up, 10/69 (14%) patients were transplanted, 26/69 (38%) died, and 16/69 (23%) patients were free from VT.

**Conclusion:** Although often a last resort, VT ablation in LVAD patients is feasible and safe when performed in experienced centers. These patients suffer from a high scar burden, and cardiomyopathy-associated rather than cannula-related scar seems to be the dominant substrate. VT recurrence after ablation is high, despite extensive treatment, and the overall prognosis of these patients is limited.

**What is known:** VT ablation in LVAD patients is one of the most complex procedures in interventional electrophysiology dealing with critically ill patients. These procedures are highly prone to technical difficulties and complications, potentially limiting procedural success and outcome.

**What the study adds:**

- Most LVAD patients requiring VT ablation have a history of ventricular arrhythmia prior to LVAD implantation, and scar-related re-entry is the predominant arrhythmogenic mechanism.
- LVAD related technical challenges are present but seem to have little impact on procedural efficacy. No association of electromagnetic interference and LVAD model was observed.
- Extensive low voltage areas were detected in most patients. Unlike in non-LVAD patients, LVAD patients showed no difference in endocardial scar between ICM and NICM.
- Neither the type of cardiomyopathy nor the endpoint of non-inducibility but the extent of myocardial scar seems to predict VT recurrence in patients with LVAD.

**Graphical abstract:** 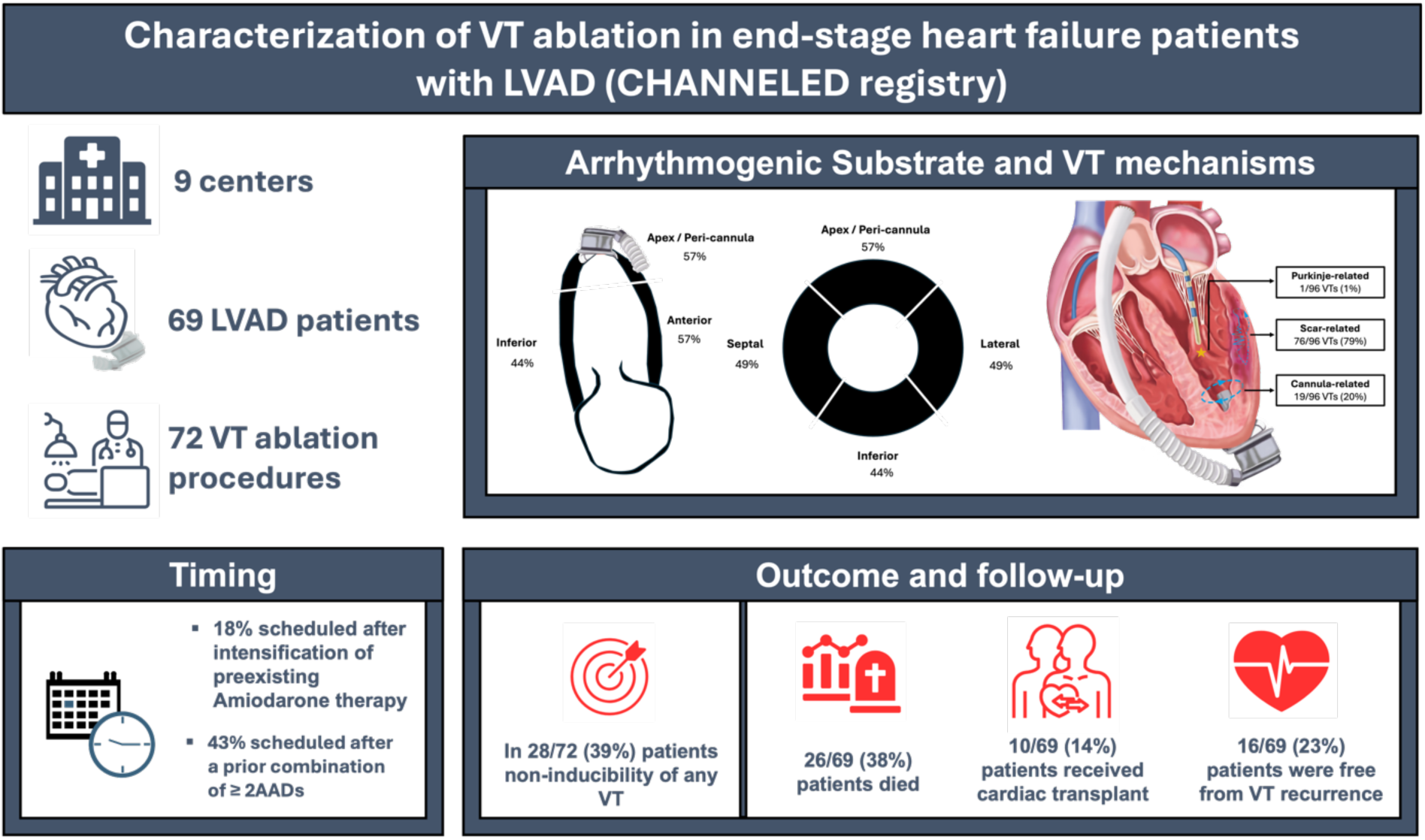

## Introduction

Left ventricular assist device (LVAD) implantation has emerged as a cornerstone therapy for selected end-stage heart failure patients, either as bridge to transplantation or as destination therapy. Expanding use of LVADs as destination therapy has led to a growing LVAD patient population [1]. Ventricular arrhythmias (VAs) are common in these end-stage heart failure patients both before and after LVAD implantation. Despite the hemodynamic support, VAs are associated with increased mortality, substantial morbidity from recurrent implantable cardioverter-defibrillator (ICD) shocks, and rapid heart failure progression especially of the unsupported right ventricle [2,3]. Regarding VA management in LVAD patients, antiarrhythmic drugs (AADs) are mostly used as first-line therapy, but as VAs become AAD-refractory there are few options other than catheter ablation (CA) [4]. Consequently, VT ablation plays an important role in the management of VAs in LVAD patients. Alongside common challenges of VT ablation in advanced heart failure patients technical and procedural challenges unique to VT ablation in LVAD carriers exist. Technical challenges such as the risk of catheter entrapment, electromagnetic interference with the surface ECG or electroanatomic mapping systems (EAM) add to the complexity of VT ablation in these patients [5,6].

Existing data on CA of VT in LVAD patients is either outdated or derived from a limited number of small case series. Hence, the extent to which these challenges impact on procedural and patients’ outcomes is not well quantified. In an era of modern LVAD devices, novel high-density EAM, and image integration availability, new data is needed to better characterize the role of VT ablation in LVAD patients. Therefore, the present multicenter registry systematically assessed substrate burden, VT mechanisms and analyzed procedural parameters and outcome of VT ablation in LVAD patients.

## Methods

This multicenter registry collected patient data from a collaboration of 9 tertiary care centers in Germany and Switzerland. All patients undergoing VT ablation after LVAD implantation in the respective study center were included retrospectively in this registry. Data were gathered from a variety of sources: Electronic medical records, detailed electrophysiological procedure reports, three-dimensional mapping systems, and ICD interrogation. Institutional review boards approved the study, with patient consent obtained as per individual center protocols. The study complies with the Declaration of Helsinki and this registry is registered on ClinicalTrials.gov (NCT06063811).

### Left ventricular assist devices

In contemporary clinical practice there are two different LVAD models available. The HeartWare ventricular assist device (HVAD™) (Medtronic Inc., Dublin, Ireland) and the HeartMate 3 (Abbott, Abbott Park, IL, USA). The HVAD™ has an impeller suspended in both, magnetic and hydrodynamic forces; although the distribution and sale of the HVAD™ System was discontinued in 2021 this third generation LVAD system is still widely present in current clinical practice [7].

The HeartMate 3 (Abbott, Abbott Park, IL, USA) device received CE mark in 2015, and FDA approval in 2017, and is a fully magnetically levitated centrifugal-flow third generation LVAD. It has shown significant survival benefit over previous axial-flow pump LVADs, e.g., the HeartMate 2 (Abbott, Abbott Park, IL, USA) and is currently the only device available for implantation [8].

### Substrate and VT mapping

CARTO3^®^ (Biosense Webster, Diamond Bar, California), EnSite™Precision or EnSite™X (Abbott, Chicago, IL, USA) was used to create substrate and/or activation maps. Regarding substrate mapping, a peak-to-peak bipolar electrogram amplitude <0.5mV was defined as dense scar, voltage ≥0.5 and <1.5mV as border zone. For the assessment of scar distribution and burden, the LV was subclassified in six different segments (basal, anterior, inferior, septal, lateral and peri-cannular) and an extensive burden was defined as the presence of low voltage in ≥ 3 segments. A multipolar mapping catheter for high-density mapping (PentaRay^®^, Biosense Webster, Inc., Diamond Bar, CA, USA or Advisor™HD-Grid, Abbott, Chicago, IL, USA) was applied at the operators’ discretion [5,9]. As an image integration technology either the CARTOUNIVU™ Module (Biosense Webster, Diamond Bar, CA, USA) was used to merge a fluoroscopy image with the electroanatomic map or a pre-procedural perfusion CT-scan was performed, which was subsequently segmented and analyzed with the inHEART system (IHU Liryc, Pessac, France). The resulting 3D-model containing information about the underlying substrate was subsequently merged with the endocardial electroanatomic map (Figure 1) [9,10]. VT mapping was defined as determination of sites for ablation if diastolic potentials were noted, a post-pacing interval within 30 ms of the VT cycle length after entrainment was observed and/or by identification of the critical isthmus after activation mapping. Substrate modification was defined as ablation of sites with low amplitude fractionated electrograms, long stimulus-to-QRS, late potentials, or the best pace map sites [5,6,9].

**Figure 1.**
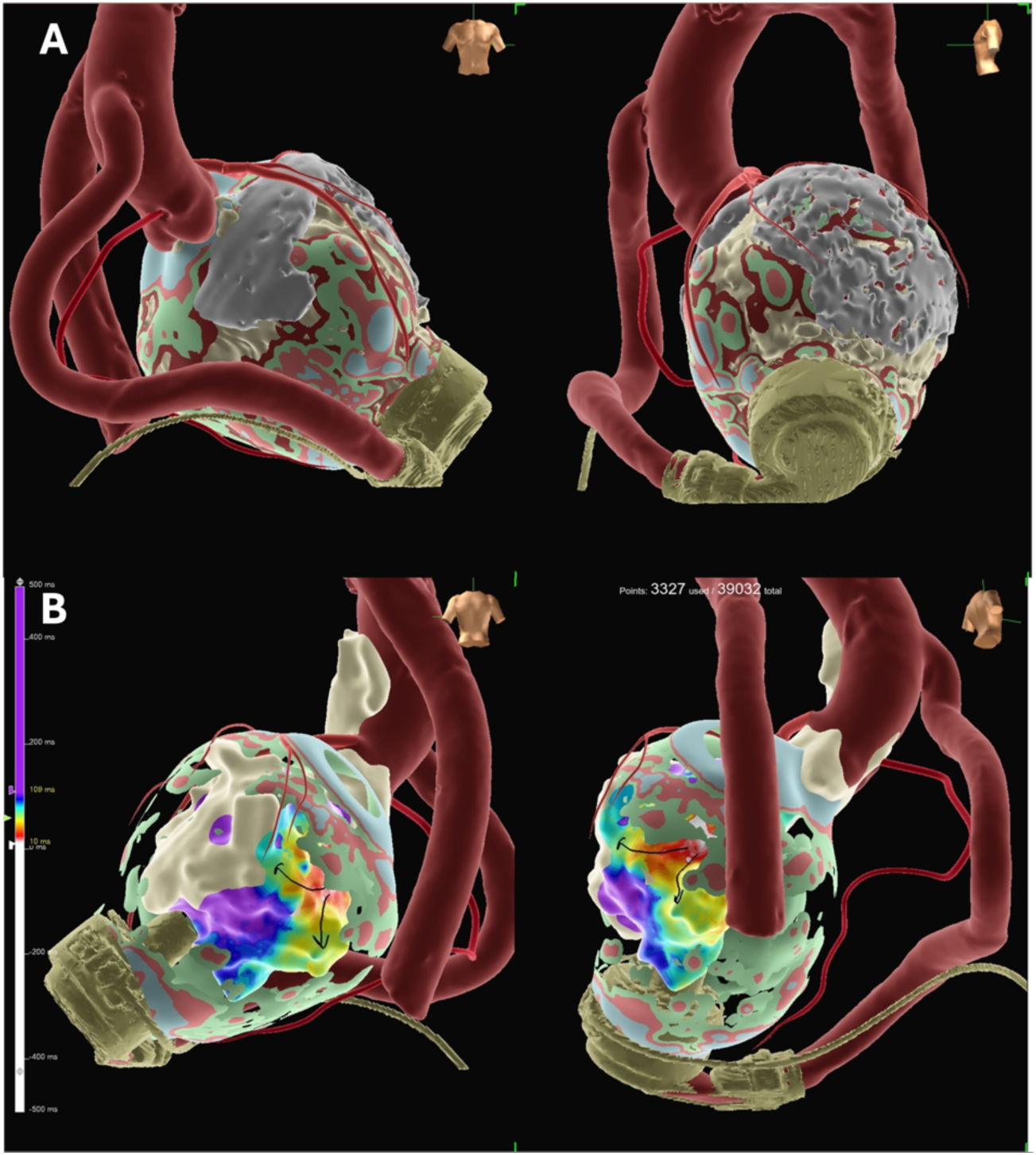
Combination of a CT-based 3D model created with Inheart and a propagation map of the clinical VT in a patient with NICM and LVAD. The 3D model shows an extensive scar burden with substrate detected at basal, septal, apical and inferior scar with a distinct basal and apical wall thinning. The model is derived from the arterial phase of the pre-procedural CT scan. The propagation map shows the clinical VT with a basal scar related reentry.

### Statistical analysis

Data analysis was performed using SPSS Statistics (Version 27, IBM, Chicago, IL, USA). Categorical variables were presented as numbers and percentages and continuous variables as mean with standard deviation or median (25th to 75th percentile) were appropriate. Categorical variables were compared between groups with chi-square tests or Fisher’s exact tests. Student’s T-tests or Mann–Whitney U tests were used to compare continuous measures between groups. ANOVA was used for multivariate analysis.

## Results

### Patient characteristics

A total of 69 patients (mean age 60.7±8.4 years, 90% male) underwent 72 VT ablation procedures. All patients had third generation LVAD devices implanted, of those 42/69 (61%) HM3 and 27/69 (39%) HVAD™ devices (Table 1). The underlying cardiomyopathy was ischemic in 41/69 (59%) patients and nonischemic in 28/69 (41%) patients with a mean left ventricular ejection fraction of 18.4%±7.1% accompanied by at least moderate right ventricular dysfunction in 33% of patients. In 33/69 patients (48%), the LVAD implantation was considered a bridge-to-transplantation, in 6/69 patients (9%) a bridge-to-candidacy, and in 30/69 patients (44%) the LVAD indication was destination therapy. An ICD was present in 63 patients (91%).

**Table 1.** Patient characteristics. BMI, body mass index; LVEF, left ventricular ejection fraction; LVAD, left ventricular assist device, ICD, implantable cardioverter defibrillator, CRT-D, cardiac resynchronization therapy defibrillator.

| Patient characteristics | N=69 |
| --- | --- |
| Male | 62 (90%) |
| Age (years) | 60.7±8.4 |
| BMI (kg/m<sup>2</sup>) | 26.9±84.9 |
| Ischemic cardiomyopathy | 41 (59%) |
| LVEF (%) | 18.4±7.1 |
| **LVAD model** |  |
| Heartmate III | 42 (61%) |
| Heartware | 27 (39%) |
| **LVAD indication** |  |
| Bridge-to-transplantation | 32 (46%) |
| Bridge-to-candidacy | 4 (6%) |
| Bridge-to-decision | 2 (3%) |
| Destination-therapy | 30 (44%) |
| **Cardiac implantable electronic devices** |  |
| Patients without defibrillator | 6 (9%) |
| Single-chamber ICD | 21 (30%) |
| Dual-chamber ICD | 12 (17%) |
| CRT-D | 30 (44%) |
| **Comorbidities** |  |
| Atrial fibrillation | 41 (59%) |
| Coronary artery disease | 46 (67%) |
| Diabetes | 17 (25%) |
| Renal failure (GFR≤50ml/h) | 23 (33%) |

### Timing and indication for VT ablation after LVAD implantation

Prior to LVAD implantation, most patients (64%) already had suffered a sustained VT and ICD shocks (Table 2). After LVAD implantation, VT occurred after a median of 279 days [42–862 days] and 19% of patients required urgent CA <30 days post implantation. An electrical storm (defined as ≥ 3 sustained VTs/ICD shocks in 24h) was common and occurred in 70% of patients. Symptoms during VT were mainly heart failure worsening (35%), LVAD alerts (17%), palpitations (17%). Less commonly syncope (8%) or lightheadedness (8%) were reported (Table 2). VT ablation was commonly scheduled after a previously escalated AAD therapy. Either after intensification of amiodarone therapy (18/72 procedures, 25%) defined as the administration of an additional loading dose of 400-600mg for two weeks or after VT recurrence despite a combination of ≥2 AADs (31/72 procedures, 43%).

**Table 2.** Characteristics and symptoms of ventricular arrhythmias in LVAD patients. VA, ventricular arrhythmia; LVAD, left ventricular assist device; ICD, implantable cardioverter defibrillator.

| Characteristics of ventricular arrhythmias | n=69 |
| --- | --- |
| **History of ventricular arrhythmia** |  |
| VT prior to LVAD implantation | 44 (64%) |
| Time from LVAD implantation to VA occurrence (days) | 279 IQR (42-862) |
| VA occurrence <30 days post LVAD implantation | 13/67 (19%) |
| Electrical storm | 35/50 (70%) |
| Number of ICD shocks post LVAD implantation | 3 IQR (1-8) |
| **VA symptoms** |  |
| No symptoms besides ICD shock | 10 (15%) |
| LVAD alarm | 12 (17%) |
| Syncope | 6 (9%) |
| Light-headedness | 6 (9%) |
| Palpitations | 11 (16%) |
| Heart failure worsening | 24 (35%) |

### Procedural characteristics

All procedures were performed at tertiary care centers with dedicated and permanent VAD team presence at the institution. Detailed procedural characteristics are presented in Table 3. Median procedure duration was 198 minutes (IQR 149-250 min.) and most procedures were performed under deep sedation (68%) while general anesthesia was used in 32% of cases. Intraprocedural vasopressor therapy was necessary in 30% of cases. An LVAD technician was present during 66% of the procedures. For LV access the preferred approach was transseptal (74%). Pericardial access was not attempted in any cases. An EAM system was used in all procedures, CARTO3^®^ in 61/72 procedures (85%) and 11/72 (15%) procedures were performed using the Ensite system (n=5 EnSite™Precision and n=6 EnSite™X). In most patients (74%) a high-density map was acquired using a multipolar mapping catheter.

**Table 3.** Procedural data. LAVA, late abnormal ventricular activation; RF, radiofrequency current; sec, seconds; min, minutes; VT, ventricular tachycardia; IQR, interquartile range.

|  |  |
| --- | --- |
| LVAD technician present during procedure | 31/47 (66%) |
| **Anesthesia** |  |
| Sedation | 49/72 (68%) |
| General anesthesia | 23/72 (32%) |
| **Mapping system** |  |
| Ensite Precision | 5/72 (7%) |
| Ensite X | 6/72 (8%) |
| CARTO | 61/72 (85%) |
| **Access** |  |
| Retrograde | 9/72 (13%) |
| Transseptal | 53/72 (74%) |
| Combined retrograde and transseptal | 10/72 (14%) |
| **VT map approach** |  |
| HD-mapping with multipolar catheter | 53/72 (74%) |
| Primarily activation map | 29/66 (44%) |
| Primarily substrate based | 37/66 (56%) |
| Entrainment mapping performed | 10/47 (21%) |
| Pace mapping performed | 28/52 (54%) |
| **VT ablation approach** |  |
| Primarily substrate-based (scar homogenization, LAVA) | 29/72 (40%) |
| Primarily ablation during VT | 3/72 (4%) |
| Combination of ablation during VT and substrate homogenization | 40/72 (56%) |
| Total number of VTs targeted | 96 |
| VTs targeted per patient | 1.9±1.8 |
| **Procedure characteristics** |  |
| Total RF-time (sec.) | 1899 IQR (1093-2798) |
| Fluoroscopy duration (min.) | 15.8 IQR (9.2-25.9) |
| Procedure duration (skin-to-skin;min.) | 197.5 (IQR 148.8-250.3) |
| **Acute outcomes** |  |
| Non-inducibility of any VT | 28/72 (39%) |
| Non-inducibility of clinical VT | 31/72 (30%) |
| Substrate-based due to prior non-inducibility | 8/72 (11%) |
| Clinical VT still inducible | 5/72 (7%) |
| **Post-procedural monitoring** |  |
| Intensive care | 35/72 (49%) |
| Intermediate care | 25/72 (35%) |
| Normal ward | 12/12 (17%) |
| Days until discharge | 5.5 IQR (3-14) |

For VT ablation either a combined approach of critical isthmus ablation and substrate homogenization was performed (56%) or, mostly due to inducibility of several VTs, a primarily substrate-based approach with scar homogenization and ablation of late potentials was carried out (40%).

Irrigated radiofrequency (RF) energy was delivered at a power of 30–65 Watts and the primary goal of all procedures was non-inducibility of any VT. Electromagnetic interference (EMI) occurred in 41% of procedures. However, in only 11% of these cases severe EMI led to incomplete map acquisition. A transient loss of catheter visualization was observed in 41%. The susceptibility to EMI was comparable between LVAD models (HM3 12/32 vs. HVAD 12/23; p=0.28). In a single case severe EMI led to abortion of the procedure. LVAD flow adaption aiming for EMI reduction was not attempted in any case.

### Scar distribution and VT characteristics

In all patients low voltage areas were detected in the endocardial voltage map and in most procedures (78%) low voltage areas were identified in more than one segment. The distribution of endocardial scar is depicted in Figure 2. An extensive scar burden was observed in 57% of patients. Of note, in most patients with non-ischemic cardiomyopathy (NICM) a high burden of endocardial scar was seen (Figure 1). There was no difference in endocardial scar burden between ischemic cardiomyopathy (ICM) and NICM patients (p=0.76) (Table 4). The predominant VT morphology was a superior axis (41%) and right bundle branch block-like morphology (68%) (Table 5).

**Figure 2.**
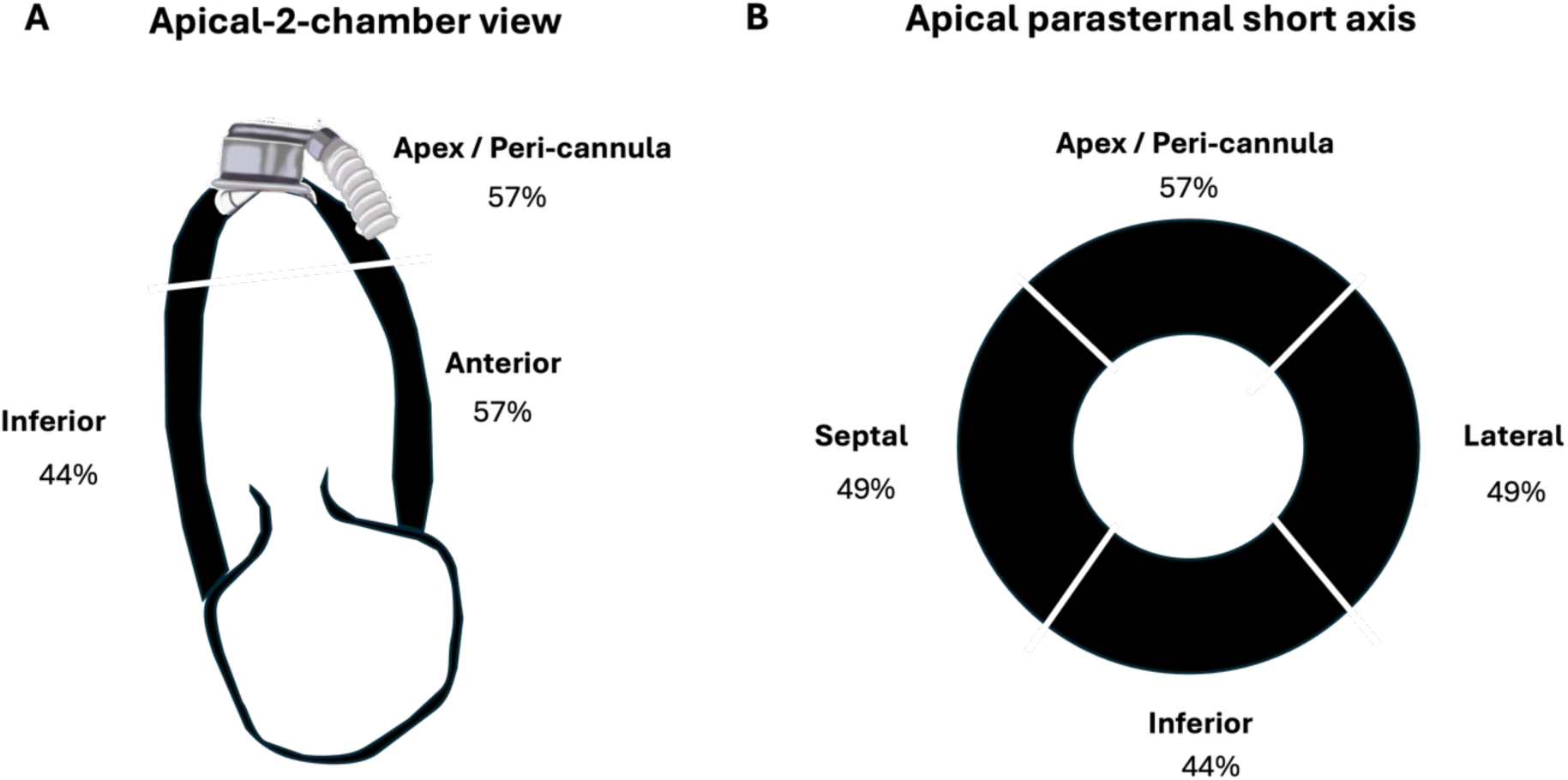
A schematic illustration of a apical-2chamber (A) and apical parasternal short axis (B) of the left ventricle. Percentages show the detection of low voltage / scar in the respective LV segment over the study cohort. At an overall high burden of endocardial scar in end-stage heart failure patients with LVAD the illustration shows a homogenous distribution of scar with all LC segments being affected comparably.

**Table 4.**
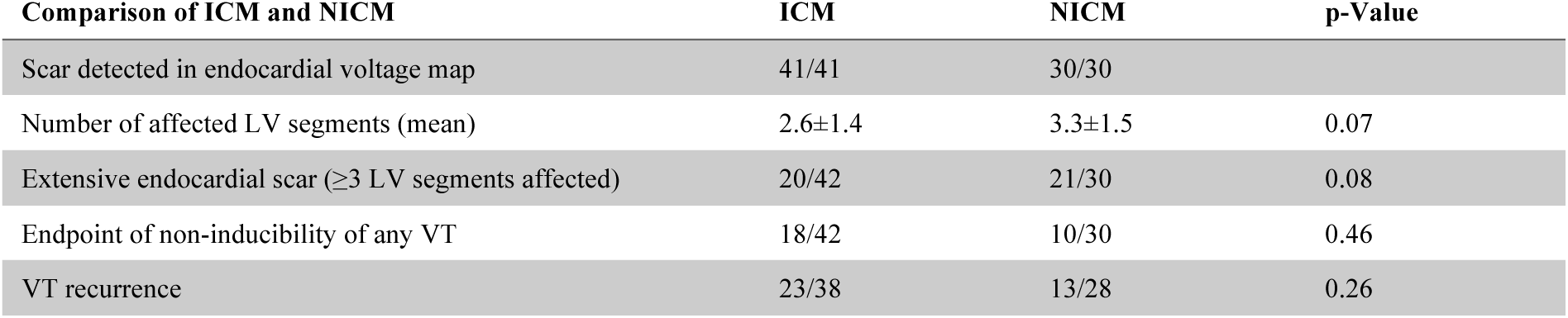
Comparison of scar burden and outcome between ICM and NICM. ICM, ischemic cardiomyopathy; NICM, non-ischemic cardiomyopathy; VT, ventricular tachycardia.

**Table 5.** ECG morphologies of VTs targeted during VT ablation in LVAD patients. ECG, electrocardiogram; VT, ventricular tachycardia, LVAD, left ventricular assist device

| VT characteristics and morphology | n=91 |
| --- | --- |
| Inferior axis (II, III, aVF positive) | 33 (36%) |
| Superior axis (II, III, aVF negative) | 37 (41%) |
| Left axis deviation (I positive, II, III negative) | 6 (7%) |
| Right axis deviation (III positive, I, II negative) | 15 (16%) |
| Left bundle branch block morphology | 15 (16%) |
| Right bundle branch block morphology | 62 (68%) |
| Negative concordance | 8 (9%) |
| Positive concordance | 6 (7%) |

Overall, 96 VTs (1.9±1.8 VTs per patient) were mapped and targeted for ablation. Table 6 shows the reported sites of successful ablation. The underlying mechanism was determined to be a scar-related in the majority of VTs (79%). Less commonly the VT mechanism was attributed to the LVAD’s inflow cannula (20%), and in one case a Purkinje-related mechanism was identified. A schematic presentation of VT mechanisms is shown in Figure 3.

**Figure 3.**
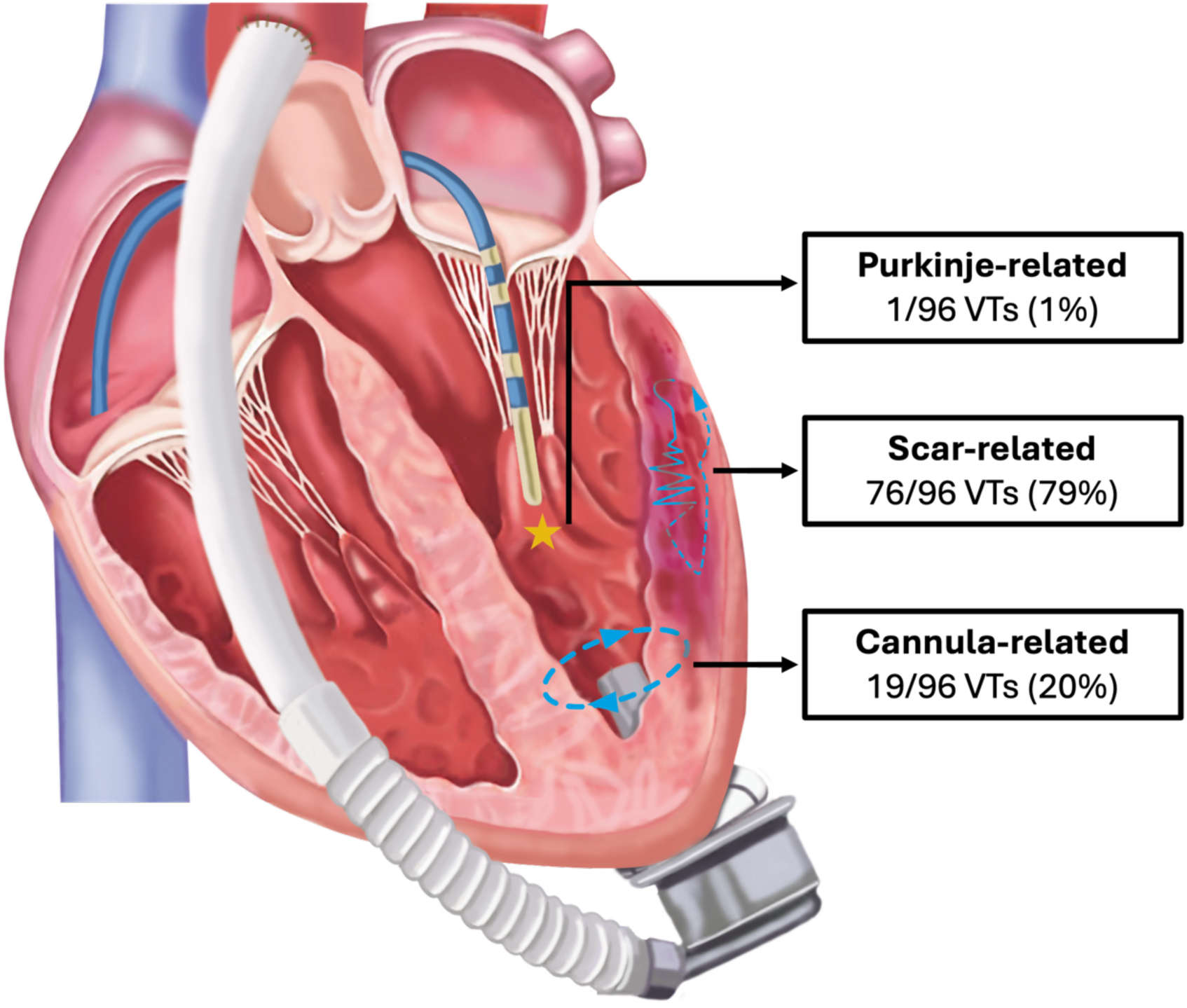
A schematic 5-chamber view of a left ventricle (LV) after LVAD implantation. An ablation catheter is introduced transeptally in the LV. The reported mechanisms of ablated VT in this end-stage heart failure patients cohort with LVAD are graphically depicted with 79% of VTs being scar related and 20% of VT were related to the cannula.

**Table 6.**
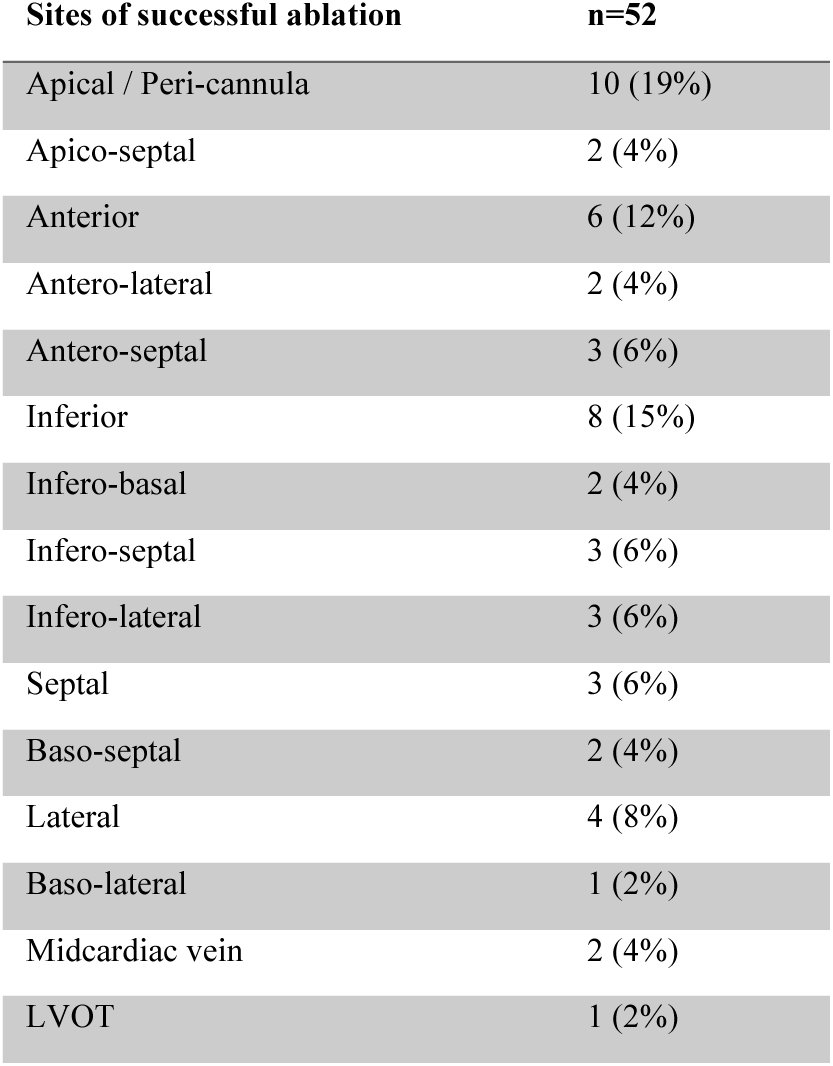
Sites of VT termination/successful ablation.

| Sites of successful ablation | n=52 |
| --- | --- |
| Apical / Peri-cannula | 10 (19%) |
| Apico-septal | 2 (4%) |
| Anterior | 6 (12%) |
| Antero-lateral | 2 (4%) |
| Antero-septal | 3 (6%) |
| Inferior | 8 (15%) |
| Infero-basal | 2 (4%) |
| Infero-septal | 3 (6%) |
| Infero-lateral | 3 (6%) |
| Septal | 3 (6%) |
| Baso-septal | 2 (4%) |
| Lateral | 4 (8%) |
| Baso-lateral | 1 (2%) |
| Midcardiac vein | 2 (4%) |
| LVOT | 1 (2%) |

### Acute outcome and discharge

The endpoint of non-inducibility of any VT was achieved in 28/72 (39%) of procedures. Acute outcomes were similar in patients with ICM and NICM (Table 4). Most patients were subsequently transferred to intensive care unit (49%) or intermediate care unit (35%) for post procedural monitoring and were discharged from the hospital after a median of 5.5 days (IQR 3-14 days). The majority was discharged with continued oral amiodarone therapy (64%) or a combined AAD therapy with amiodarone and mexiletine (21%).

### Procedural complications

Procedure related complications were reported for 9/72 procedures (13%). Of those, the majority were access related (6/9, 67%). One pericardial effusion occurred, one iatrogenic aortic dissection between the right iliac artery and the renal branches occurred. One procedure-related death (1.4%) was reported: The patient suffered from a post-procedural oxygen desaturation; despite immediate intubation the patient died later from hypoxic brain injury. No LVAD associated complications were reported.

### Long-term outcome and mortality

Follow-up information was available for 66/69 patients, with a median follow-up of 283 days (IQR 70-587 days). Recurrence of VT occurred in 36/66 (55%) of patients. There was no difference in VT recurrence rates (figure 4) between NICM and ICM patients (NICM 13/28, 46% vs. ICM 23/38, 61%; logrank p=0.19). The number of low voltage areas and consequently the extension of left ventricular scar was associated with VT recurrence (ANOVA p=0.03; r^2^=0.1), whereas the procedural endpoint of non-inducibility of any VT did not predict VT-free survival (ANOVA p=0.22). During follow-up 10/66 patients (15%) received a cardiac transplant after a median of 533 (IQR 243-866) days after the VT ablation procedure. Overall, 26/66 patients (39%) deceased. Of these, 9 patients (14%) died during the first 30 days after VT ablation and a further 17/26 patients deceased during follow-up after a median time of 195 days (IQR 83-506 days) primarily from heart failure, LVAD failure, sepsis, or intracranial bleeding. None of the deaths was directly attributed to VA, but one patient died from procedure associated hypoxic brain damage as a result of postprocedural hypoxemia.

**Figure 4.**
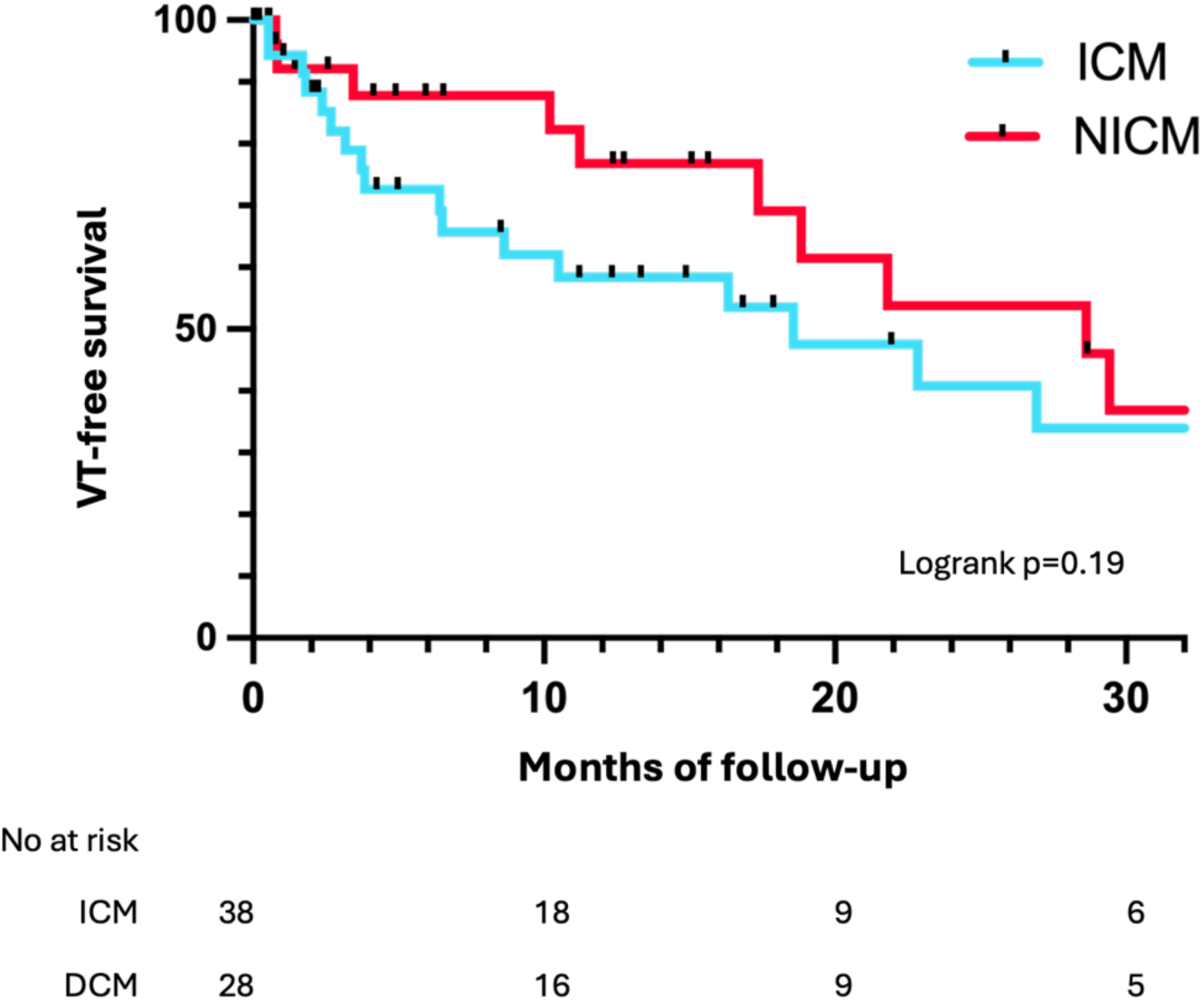
Kaplan-Meier analysis of VT-free survival after VT ablation. ICM, ischemic cardiomyopathy; NICM, non-ischemic cardiomyopathy.

## Discussion

VT ablation in end-stage heart failure patients equipped with an LVAD is among the most complex and challenging procedures in interventional electrophysiology. Currently available data are mostly outdated or derived from small case series [5,6]. This multicenter analysis represents the largest cohort of patients with third generation LVADs undergoing VT ablation, and provides several important findings:

1. Catheter ablation of VT in LVAD patients is feasible and can be performed safely in experienced centers but 30-day mortality was 14% in this critically ill patient cohort. LVAD related technical challenges were present but had little impact on procedural efficacy. Importantly, no association of EMI and LVAD model was observed.
2. VT recurrences occur in over 50% of patients during long-term follow-up.
3. Outcome and recurrence rates were comparable between NICM and ICM, and VT recurrence remains high despite maximal therapy. Not procedural endpoints or the underlying cardiomyopathy, but the extent of endocardial scar was associated with VT recurrence.
4. Most LVAD patients requiring VA treatment had a history of VA prior to LVAD implantation, and scar-related re-entry was the predominant arrhythmogenic mechanism.
5. An overall high burden of endocardial low voltage was detected. Unlike in non-LVAD patients, in LVAD patients no difference in endocardial scar between ICM and NICM was observable.

### Safety and technical aspects

The rate of major complications was low, and procedure associated complications were mostly attributed to the vascular access site. No LVAD-associated complication or catheter entrapment were observed. The overall rate of procedural complications (13%) in this cohort was comparable to non–LVAD VT cohorts of patients with structural heart disease [11], suggesting a reasonable safety profile for VT ablation in LVAD patients.

30-day mortality was 14% and most deaths occurred during the initial hospitalization after VT ablation. Of those, one procedure related death was reported but most patients died from terminal heart failure (7/9 patients, 78%) or non-procedure related septic shock (1/9 patients) and LVAD-failure (1/9 patients) underlining the critical condition of these patients.

Regarding the technical challenges of LVAD VT ablation, several issues including EMI affecting surface-ECG and mapping system, transient loss of catheter visualization, hindered assessment of catheter proximity to the inflow cannula, or incomplete map acquisition have been described and discussed previously [6,9,12]. Specifically, concern had been raised regarding the HM3 system with its fully magnetically levitated impeller being particularly prone to EMI with potentially critical impact on VT ablation [9]. Present cohort of VT ablation in third generation LVADs indeed confirmed previously described EMI, including transient loss of catheter visualization, but only minimal impact on procedure efficacy was reported. Regarding the occurrence of EMI with different LVAD systems, no difference in EMI severity was observed between the HVAD and HM3 system. Interestingly, although EMI was observed, in none of the cases LVAD flow or turbine speed were modulated, and not every procedure had an LVAD technician present.

### Acute and long-term outcome

Especially given the patients’ clinical condition, the advanced disease state and complexity of the technical setup, VT ablation in LVAD patients was associated with a satisfactory acute outcome with 82% of patients being either non-inducible for any VT or the clinical VT. This is comparable to previously published outcomes of VT ablation in LVAD and non-LVAD patients [6,13]. Nevertheless, recurrence of VT remains frequent and occurs in over 50% of VT ablations in these patients (figure 4). Similar to previously published smaller cohorts, the procedural endpoint of non-inducibility did not predict VT-free survival after VT ablation in LVAD patients [9,14]. However, the stage of disease seems to impact VT-free survival. In this analysis, the extent of LV scar was associated with VT recurrence. It can be hypothesized that further heart failure deterioration over time creates further substrate resulting in arrhythmia recurrence irrespective of the procedural endpoint. Regarding long-term outcome, the overall prognosis remains poor with a mortality of 39% during follow-up despite maximal therapy. This is in line with outcome data from LVAD cohorts in the absence of VT ablation [15].

### VT mechanisms, scar burden and implications for outcome

In the past, multiple causal mechanisms have been suggested for VA in LVAD patients. Foremost, the addition of an arrhythmogenic substrate related to the incision necessary for inflow cannula placement may substantially contribute to post LVAD VA [6,16]. Consequently, there are several arrhythmogenic mechanisms after LVAD implantation, potentially requiring different therapeutic approaches. The available literature [6,9] indicates that besides new potential arrhythmogenic mechanisms, most VTs are scar related re-entries and this study strongly supports this observation: Most LVAD patients requiring VA treatment had a history of VA and ICD shocks prior to LVAD implantation indicating the underlying substrate as being the primary cause. Scar-related re-entry was identified as the predominant arrhythmogenic mechanism (79% of VTs) and the extent of endocardial scar was associated with VT recurrence.

Irrespective of the underlying cardiomyopathy, we found a high burden of endocardial low voltage, which can be interpreted as an electroanatomic correlation of the histological findings of Strecker et al. [17] indicating severely altered myocardium in end-stage heart failure patients with LVAD. Although the LVAD implantation itself does contribute to arrhythmogenesis, in consideration of these findings and previously published data it is reasonable to assume, that VA and adverse outcomes of VT ablation in patients with LVAD are mostly attributable to end-stage heart failure itself and the disease progression.

### Future considerations for VT ablation in LVAD patients

VT ablation in end-stage heart failure patients with LVAD is a complex procedure requiring the collaboration of heart failure specialists, LVAD technicians, and electrophysiologists. These procedures should therefore only be performed in specialized centers. In this setting, such procedures have shown a satisfactory safety profile and acute outcome. Nevertheless, technical challenges, VT recurrence and mortality remain high. Based on present findings and current literature the following considerations may further improve the workflow and the outcome of VT ablation in LVAD patients: Catheter ablation of VT in LVAD patients is often chosen as a last resort, after multiple ICD shocks, electrical storm, or recurrent ICD shocks besides a combination of ≥ 2 AADs [18]. Given the high overall mortality, the impact of ICD shocks on quality of life and mortality, the reasonable safety profile with satisfactory acute outcomes and VT free survival, especially considering recently published data showing improved survival of LVAD patients undergoing VT ablation compared to medical therapy alone [19], VT ablation should be considered earlier in these patients.

During VT ablation, electromagnetic interference between the LVAD and the EAM and surface ECG is a common phenomenon. Although EMI severely affected procedural outcome in only a limited number of patients in our study, there is still room for improvement.

Reducing the LVADs’ rotor speed may reduce EMI. Especially mapping and ablating in proximity of the inflow cannula remains challenging. For further improvement advanced image integration techniques and integration of CT-based 3D models may further facilitate catheter movement and potentially overcome catheter visualization issues in the vicinity of the cannula (Figure 1). Regarding VT mechanisms and arrhythmogenic substrate in LVAD patients with severely affected myocardium, the demarcations between the underlying cardiomyopathies appear to be indistinct. Neither the type of cardiomyopathy nor the endpoint of non-inducibility but the extent of myocardial scar seem to predict VT recurrence and septal scar was previously identified as a predictor for VT recurrence in LVAD patients [9]. The extent of substrate and the severity of heart failure seem to limit procedural success and patient’s outcome. In this context a distinct substrate evaluation, high-density mapping, pre-procedural imaging guided substrate evaluation, alongside with an even more extensive substrate modification approach and application of bipolar ablation (e.g. in septal scar) may improve procedural success and should be evaluated further.

Whether VT ablation prior to LVAD implantation or surgical ablation during LVAD implantation may increase efficacy and decrease VT occurrence during follow-up remains unclear. This needs to be carefully considered as for example epicardial ablations are usually not feasible after LVAD implantations.

## Limitations

Present study is a multi-center, non-randomized retrospective registry with the inherent limitations. As a multi-center registry, it includes different procedural approaches, mapping, and ablation strategies. However, it reflects the current practice in 9 tertiary care centers. Although present registry provides the largest cohort of LVAD patients undergoing VT ablation, given this specific subset of patients studied, the overall sample size remains small. This precludes advanced statistical analysis. Furthermore, all considerations regarding future improvements of VA treatment in LVAD patients should be viewed as explorative and hypothesis generating. Prospective randomized trials are needed to determine the optimal timing of VT ablation, optimal ablation strategies, endpoints, and impact of VT ablation on quality of life and mortality in end-stage heart failure patients with LVAD.

## Conclusion

Catheter ablation of VT in LVAD patients is feasible but should be performed only in experienced centers due to its complexity. Irrespective of the underlying cardiomyopathy, LVAD patients have a high burden of endocardial scar. Intrinsic myocardial scar rather than LVAD associated factors seems to be the dominant arrhythmogenic mechanism. The extent of endocardial scar was associated with VT recurrence. The recurrence of VT after ablation remains high despite maximal therapeutic efforts and the prognosis of these patients is poor.

## Non-standard abbreviations and acronyms

AAD: antiarrhythmic drugs
CA: catheter ablation
EAM: electroanatomic mapping system
EMI: electromagnetic interference
ICD: implantable cardioverter-defibrillator
ICM: ischemic cardiomyopathy
LVAD: left ventricular assist device
NICM: non-ischemic cardiomyopathy
RF: radiofrequency energy
VA: ventricular arrhythmia
VT: ventricular tachycardia

## Data availability

Data are available upon reasonable request made to the corresponding author.

## Supplemental material

Tables S1-S5

